# The Middle-Out Perspective – an approach to formalise ‘normal practice’ in public health advocacy

**DOI:** 10.1101/2021.11.17.21266405

**Authors:** Jennifer S Mindell, Yael Parag, Suzanne E. Bartington, Laura Stoll, James Barlow, Kathryn B. Janda

## Abstract

**Background:** The Middle-Out Perspective (MOP) provides a lens to examine how actors positioned between government (top) and individuals (bottom) act to promote broader societal changes from the middle-out (rather than the top-down or bottom-up). The MOP has been used in recent years in the fields of energy, climate change, and development studies. Public health practitioners involved with advocacy activities and creating alliances to amplify health promotion actions will be familiar with the general MOP concept if not the formal name.

**Methods:** This paper introduces the MOP conceptual framework and customises it for a public health audience by positioning it among existing concepts and theories for actions within public health. Using two UK case studies (increasing signalised crossing times for pedestrians and the campaign for smoke-free legislation), we illustrate who middle actors are and what they can do to result in better public health outcomes.

**Results:** These cases studies show that involving a wider range of middle actors, including those not traditionally involved in improving the public’s health, can broaden the range and reach of organisations and individuals involving in advocating for public health measures. They also demonstrate that middle actors are not neutral. They can be recruited to improve public health outcomes, but they may also be exploited by commercial interests to block healthy policies or even promote a health-diminishing agenda.

**Conclusions:** Using the MOP as a formal approach can help public health organisations and practitioners consider potential ‘allies’ from outside traditional health-related bodies or professions. Formal mapping can expand the range of who are considered potential middle actors for a particular public health issue. By applying the MOP, public health organisations and staff can enlist the additional leverage that is brought to bear by involving additional middle actors in improving the public’s health.

**Graphical abstract:** 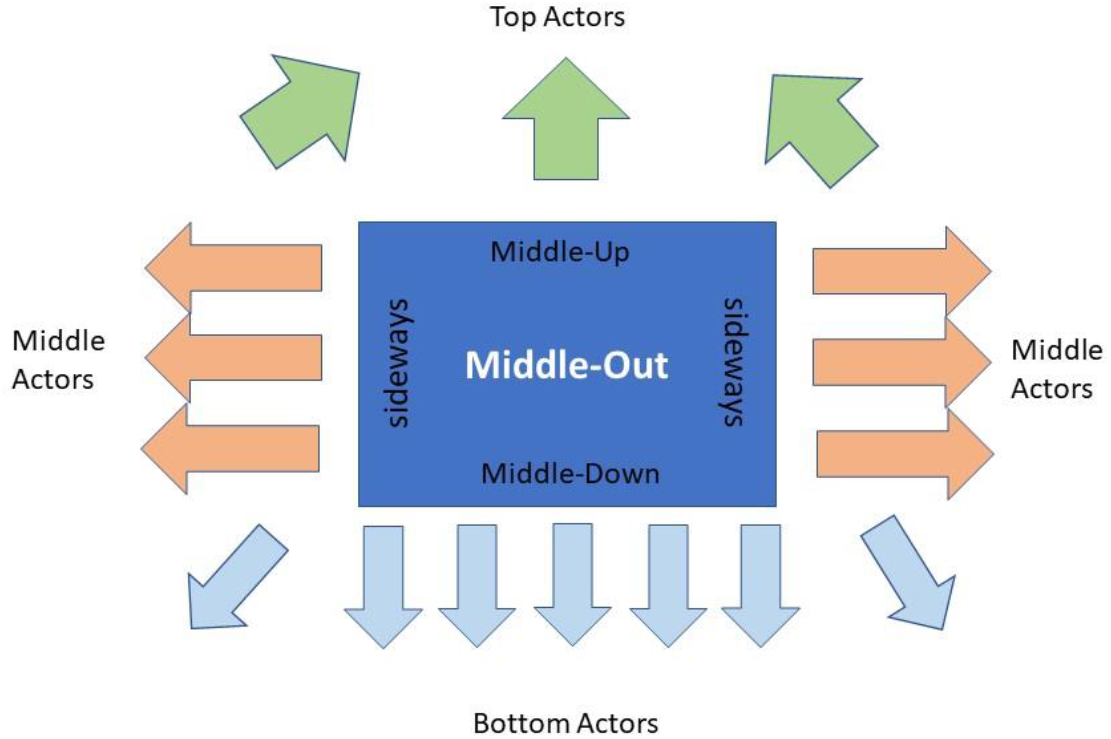

## INTRODUCTION

Both the general public and professionals in a wide range of disciplines are familiar with ‘top-down’ and ‘bottom-up’ approaches for mobilising change. Top-down actions use governments’ regulatory powers and fiscal influences to introduce or impose policy measures, such as controlling the Covid-19 pandemic. Bottom-up activities involve grassroot and individual actions (including purchasing power) to change groups’ or institutions’ behaviours at a local, regional or national level: the #MeToo and the Black Lives Matter movements are recent international examples.

Public health practitioners and organisations are generally positioned between national government and the general public. They work by assembling, reviewing and disseminating evidence and trying to influence upwards to government and downwards to local communities and individuals. However, this model underplays an essential component of effective public health working: liaising with, influencing or supporting others who are also in ‘the middle’.

The ‘Middle-Out Perspective’ (MOP) is a socio-technical framework first described by Janda and Parag in 2013 ^1^. They showed how various groups of actors positioned between actors at the top and the bottom, i.e., middle actors such as public health practitioners and other organisations working to improve the public’s health, exert their influence in three directions: middle-up, middle-down, and sideways (Figure 1). They also examined the modes by which influence was exerted: ‘enabling’, ‘mediating’ and ‘aggregating’.^1^ This approach was described initially in the field of energy and the transition to low carbon systems.^2–4^ Kranzler *et al*. applied the MOP in the field of public health to identify and focus attention on stakeholders positioned between the policy-makers generally associated with ‘top-down’ approaches and those involved with ‘bottom-up’ actions, calling for MOP to be incorporated into the public health skill set.^5^

**Figure 1.**
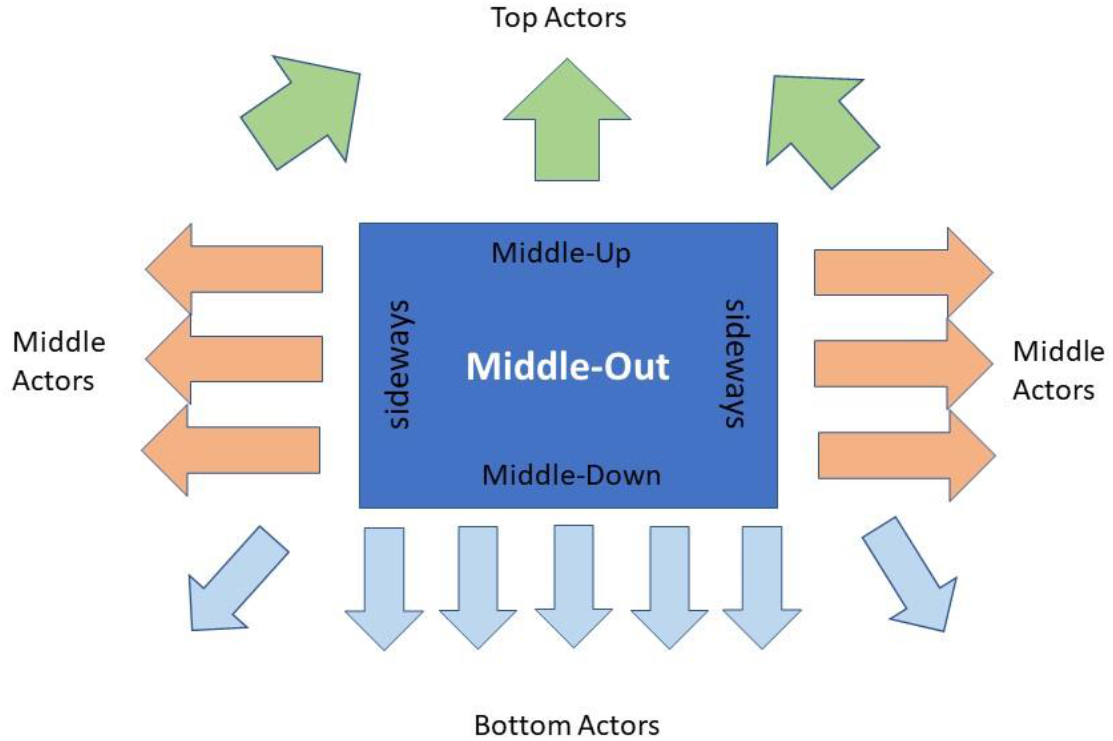
The Middle-Out Perspective.

### Existing concepts and theories for public health actions

Public health has a tradition of integrative leadership and advocacy, including coordination of individuals, organisations and communities with diverse perspectives to bring about concerted actions for equitable population health benefit.^6^ At the core of public health practice is addressing the ‘wider determinants’ of health, a diverse range of social, environmental and economic conditions and commercial influences^7–10^ which impact upon physical and mental health and contribute to health inequity.^11^ The mechanisms by which such factors influence health are dynamic and inter-related, involving a diverse array of multi-sectoral stakeholders operating within a broad, complex system, which the public health community must effectively navigate and ultimately influence to achieve desired outcomes. Therefore, public health professionals are well accustomed to operating beyond organisational ‘silos’. Yet the role of critical actors who are in the ‘middle’ of the system is often neglected in traditional public health practice.^12^ Existing conceptual models include characterisation of preventative public health action reflecting targeted interventions for ‘upstream’ health determinants (structural, affecting the population) and ‘downstream’ (individual) minimisation of harmful consequences,^13^ and application of systematic methodological frameworks, e.g., Health Impact Assessment processes.^14^

Nearly all health promotion programmes and public health policy initiatives involve changes in people’s behaviour and practices and the introduction of new norms and procedures. Their success depends on multi-faceted efforts, requiring collective action to tackle and overcome different societal, technological and economic challenges. Thus, actors such as government and regulators collaborate with public, third sector, and sometimes private organisations and the public to achieve goals. In other words, actors positioned at the top, bottom and middle change the way public health programmes are developed, implemented and regulated.^5^

Although public health research and evaluation have traditionally adopted linear cause and effect models, the complexity of public health systems^15^ and interventions^16^ are increasingly recognised.

Health in All Policies (HiAP) is an established conceptual public health approach, which seeks synergies in cross-sector actions to improve population health and equity.^17^ A HiAP approach inherently encompasses a broad spectrum of activities, from single collaborations with individual policymakers to ongoing multi-agency collaborative processes, with diverse stakeholders, including those who do not consider themselves as operating within the public health sphere.^11^ Such advocacy comprises three pillars: information, strategy, and action,^18^ requiring multiple participant roles and levels of engagement and involvement across the information, strategy and action domains. While recognising public health professionals’ direct advocacy role, this framework does not explicitly recognise the key role of additional relevant actors, both individual and organisational, and the influence of potential ‘middle actors’ including those not traditionally considered public health actors, e.g. builders.^3^ Similarly, existing research on public health advocacy has a narrow focus, typically considering health message articulation and communication within the professional or practitioner community.^7,19,20^

The MOP conceptual framework focuses on middle actors and examines how they can promote (or diminish) action by enhancing top, bottom and other middle actors’ interest in action and ability to act. In the public health field, middle actors include a wide variety of organisations that can contribute substantially to making a case for a new or amended policy or its successful implementation, to improve health and reduce inequalities. They include local government (policymakers and practitioners); higher education institutions; third sector organisations; community, interest, or industry groups; private businesses; religious organisations; and professional associations. We propose the MOP to address the existing over-simplification of ‘top-down’ and ‘bottom-up’ approaches by providing a lens through which to view public health advocacy work and identify other actors and activities that can be recruited to progress a public health agenda. Such processes thereby acknowledge the contribution of middle sector actors beyond the core professional public health community. In this paper we describe the MOP and analyse two case studies through the MOP lens.

## METHODS

### Theoretical basis of the MOP

#### Middle actors

Parag and Janda identified specific attributes necessary to be considered as middle actors (Box 1).^1,4^ Kranzler and colleagues described the domain of middle actors as *“elusive administrative spaces”* within which they *“shape policies, steer funding and facilitate continuity”*.^5^ Through these domains and activities, middle actors can exert their influence, upwards to policy makers, downwards to the public, and sideways on other middle actors in the policy arena.

Middle actors can be the immediate target that public health is aiming to influence because of their potential to be powerful allies or communication channels for knowledge exchange and suggested actions. They may be entities that affect the public’s health, without being recognised as public health organisations, such as companies providing public transport information.

**Box 1. Attributes of middle actors**

- Institutions that are visible, separate and specified, e.g. through:
  - organisational structures
  - membership
  - procedures or rules, whether official or not
- Have access to:
  - unique resources, for example
    ▪ funding
    ▪ equipment
  - other resources, for example
    ▪ expertise
    ▪ experience
- Have a distinct authority and legitimacy:
  - Professional, legal and rational
  - Spiritual and ethical
  - Traditional and charismatic
- Have pre-existing formal and/or informal channels of communication with:
  - their own members
  - other middle actors
  - top actors (e.g., decision-makers, policymakers)
  - bottom actors (e.g., individuals and citizens)

*Expanded and adapted from Janda and Parag*^1^ *and Parag and Janda*^4^

#### How middle actors influence others

Tackling complex public health challenges requires the adoption of complex and multi-disciplinary interventions that take account of contexts, actors, and environments. In such a turbulent and ‘messy’ arena, middle actors are important. How do middle actors contribute to long lasting and sustainable programmes and policies? The main mechanisms identified by Janda and Parag^1,4^ are *mediating, aggregating*, and *enabling*, although these sometimes overlap.

Middle actors act as *mediators* between the various actors in the field, often functioning as an effective communication channel, and as translators of needs and limitations. They *aggregate* various resources, e.g. knowledge and funding, to make them more robust and visible to the other actors in the field. They use their own unique resources and legitimacy to *enable* action by removing or overcoming different types of contextual, technical, normative barriers and obstacles.^1^ These modes of action occur both within public health^5^ and elsewhere.^2^

*Mediating* is particularly suited to public health practitioners’ strengths in using language appropriate for different audiences and, where necessary, ‘interpreting’ between different professional or disciplinary groups, policy-makers, and the public, including giving a voice to those frequently under-represented in research and policy debates.^19^ Health practitioners are positioned well to *aggregate* fragmented evidence and local knowledge into a comprehensive, robust and trustworthy reflection of the field. The aggregation makes scattered phenomena visible to other actors in the field. They can also aggregate relatively small budgets from different sources into a more meaningful amount, supporting more substantial action. Their unique resources that other actors lack, including moral, professional and ethical legitimacies and access to tacit and local knowledge, help them overcome barriers for change and *enable* (or delay or block) action.

#### Case studies

We selected two case studies in which non-governmental and public health organisations (some traditionally involved in health promotion and some not) have worked collaboratively to achieve national policy changes.

##### Case study 1. Smokefree legislation

Successive governments in England have had a longstanding commitment to voluntary agreements with industry for tobacco control^21^ and other public health areas. The legislation banned smoking in indoor public places, including workplaces, places of entertainment, shops, transport, etc., and reinforced existing local initiatives on public transport, for example.

##### Case study 2. Signalised pedestrian crossings

Pelican signalised pedestrian crossings (Figure 2a) have two pedestrian phases. The ‘invitation to cross’ (the ‘green person’ showing), lasts 6-10 seconds in the UK, dependent on road width. This is followed by the ‘clearance time’ (a flashing green icon or nothing visible to pedestrians), so those who are already crossing the road can reach the other side before the road traffic resumes. The clearance time duration assumes a walking speed of ≥1.2m/s (4.3km/hr, 2.7mph) in the UK; the limited time available deters mobility, rather than causing injuries.

**Figure 2.**
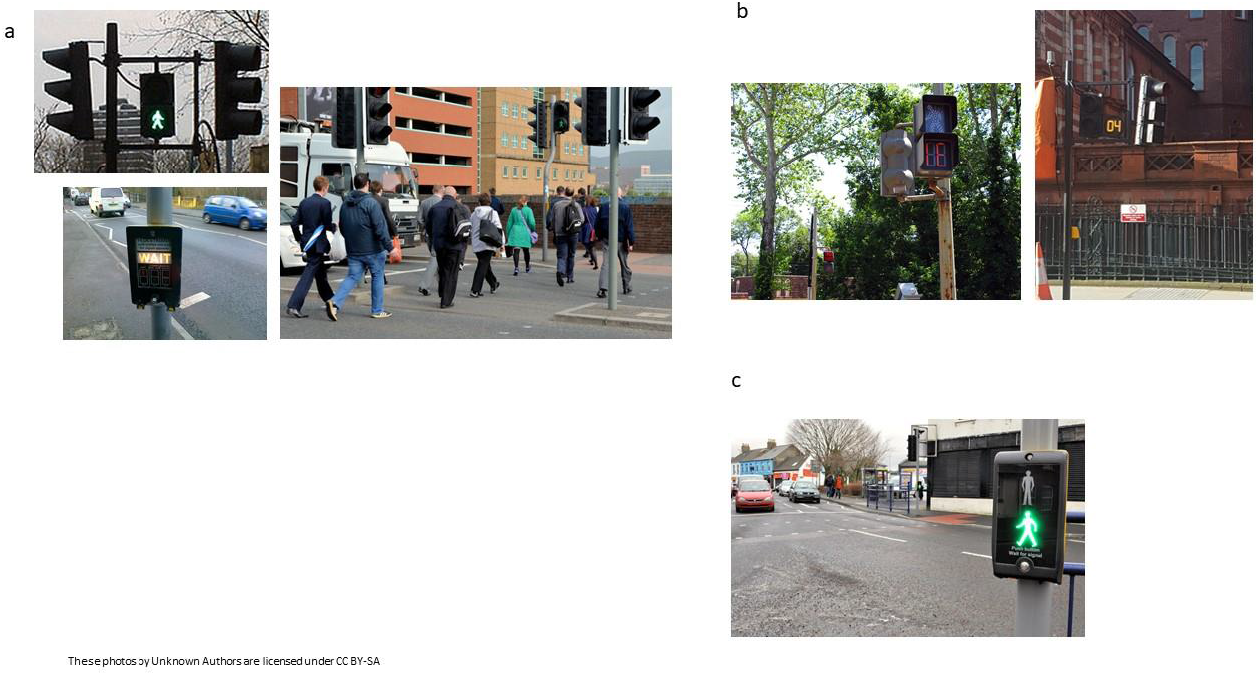
Types of signalised pedestrian crossings in the UK. Footnote: 2a: Pelican crossing (fixed timings); 2b: Countdown crossing (fixed timings); 2c: PUFFIN crossing (camera-controlled).

The clearance time duration assumed an average walking speed for the general public but did not take account of slower walking speeds for the elderly. Due to pressure initiated by an NGO, the signal crossing time was changed.

For each case study, we analysed who the actors were and how the actions taken by the key actors were used, applying the MOP framework described above.

## RESULTS

### Case study 1: Smoke-free legislation in England

An account of the advocacy work by a consortium of NGOs and practitioners’ organisations that led to the national government in England passing smoke-free legislation in 2006, implemented in 2007, has been published elsewhere.^23^ The top, middle and bottom actors are shown in Figure 3. Bottom actors were very wide-ranging in their backgrounds, knowledge of the issue, and concerns. The tobacco industry also used a middle-out approach, working through front organisations and hospitality trade associations, encouraging them to recruit their own bottom actors to lobby government to oppose smoke-free legislation (see below).

**Figure 3.**
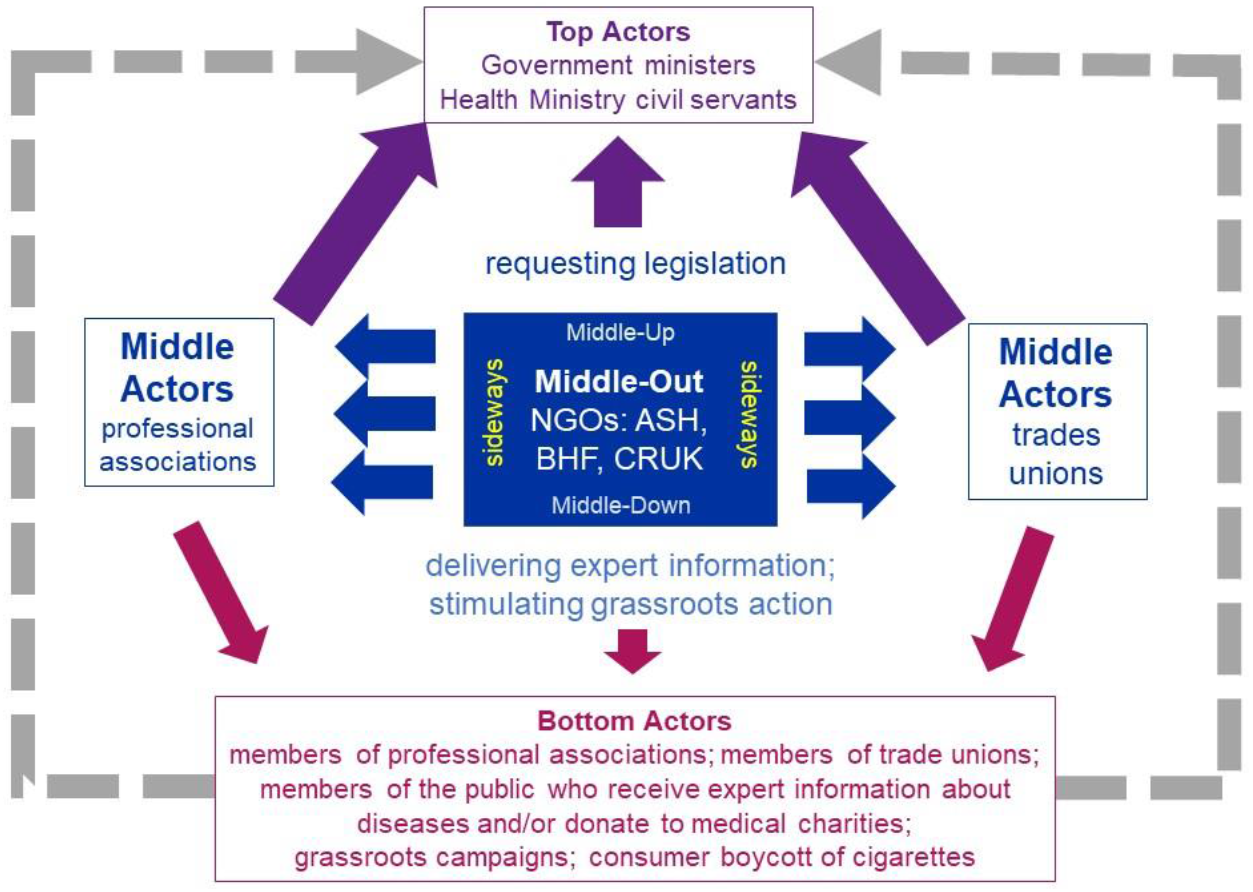
The Middle-Out Perspective used in advocating for smoke-free legislation.

The process was led by the NGO Action on Smoking and Health (ASH, www.ash.org.uk). The initial action was to build a coalition to advocate for smoke-free legislation to protect children and non-smokers from secondhand smoke. ASH was closely familiar with the action and interests of many other middle actors, and with decision-makers’ attitudes and pressure put on top actors preventing them from adopting new actions on smoking. ASH understood that while many small organisations advocate action against smoking, they can be invisible to decision-makers and their voice is not heeded. *Aggregating* these voices in a coalition made them more visible and their demand more influential. ASH’s professional expertise and reputation made them a trustworthy actor and granted them a professional legitimacy in the tobacco policy domain. The middle actors’ coalition’s activities are summarised in Table 1.

**Table 1.** Smoke-free legislation in England.

| **Direction of activity** | **Enabling<sup>a</sup>** | **Mediating<sup>a</sup>** | **Aggregating<sup>a</sup>** |
| --- | --- | --- | --- |
| Middle-down | *Using professional legitimacy to build support:* Using ASH's reputation for impartial evidence to educate the public via the media. | *Communication with the public* to increase interest and motivation: Middle actors generated grassroots support, framing the issue around improving the public's health, reducing disease, and protecting employees from occupational exposure to a lethal substance.<br>*Translating between professional and public languages:* ASH filled the role of mediators between the scientific knowledge and evidence on the relations between smoking and health, publishing reports to maintain interest and expand knowledge. Middle actors built up public | *Development of public knowledge of the risks of secondhand smoke and public support,* to improve its interest to engage in the debate through communicating with the public via the media and with members/supporters<br>*Formalisation of a grassroots initiative:* Middle actors encouraged their supporters to write to their MPs to garner |

| Direction of activity | Enabling<sup>a</sup> | Mediating<sup>a</sup> | Aggregating<sup>a</sup> |
| --- | --- | --- | --- |
|  |  | knowledge of the risks of SHS support by tailoring their messages to their members' particular focus. | political support, and demonstrate to national government that there was widespread public support. |
| Middle-up | *Building relationships:* ASH and other middle actors developed relationships with key individuals advising government<br>*Lobbying:* Middle actors lobbied government directly; and wrote to MPs and Peers.<br>*Influencing the drafting of legislation:* Legislation proposed by ASH and its | *Translating between professional and sectoral languages:* ASH filled the role of mediators between the scientific knowledge and evidence on the relations between smoking and health.<br>They proposed an evidence-based agenda as an alternative to the one proposed by the tobacco industry<br>*Building political pressure:* Middle actors were able to demonstrate to national government that | *Aggregating the evidence from the field* to present top actors with a comprehensive and more complete view of the issue:<br>Middle actors aggregated the evidence, which supported their position. |
|  | coalition of middle actors was supported in a report by the cross-party House of Commons Health Select Committee. Many of the experts who gave evidence to the Select Committee were themselves middle actors. | there was widespread public support. Core middle actors responded immediately with a letter published in a leading national newspaper, signed by senior figures from FPH, CIEH, the national public health association, and others, decrying the Secretary of State [Minister] for Health's comments that poor women needed to smoke. |  |
| Sideways | *Identifying legal/financial levers:*<br>Middle actors worked with employers, employees, and lawyers to raise the threat of legal action by employees. | *Building and maintaining coalitions with allies:*<br>ASH's initial sideways work was with an existing core group of middle actor organisations.<br>*Expanding the coalition:* Middle actors worked within their own localities to generate political | *Aggregating the evidence:*<br>Middle actors used evidence to support their position with other hospitality trade middle actors. |
|  | *Identifying new policy levers:* Middle actors used the threat of local legislation in Liverpool and London to divide the hospitality trade from the tobacco industry. | support within local government authorities, which then became additional middle actors.<br>ASH and CIEH recruited local authorities. Fifty stated they were interested in becoming smoke-free; some went further.<br>*Translating between professional and sectoral languages:* ASH filled the role of mediator between the scientific knowledge and evidence on the relations between smoking and health. | ASH supported the London Health Commission's consultation 'The Big Smoke Debate', aiding publicity and encouraging grassroots and middle actors to respond. Six other regions followed suit, broadening the extent of local dissemination of the evidence. |

<sup>a</sup> Enabling: enabling action by using own resources and legitimacy to overcome barriers; Mediating: providing a communications channel;
Aggregating: providing resources, e.g. knowledge, funding.
ASH: Action on Smoking and Health; CIEH: Chartered Institute for Environmental Health; FPH: UK Faculty of Public Health of the Royal Colleges
of Physicians

However, the tobacco industry is also a powerful middle actor active in the smoking policy arena, driven by a strong economic incentive not to limit smoking. The tobacco industry used the hospitality industry as their own middle actors, working sideways to encourage vociferous opposition to the proposed legislation by clubs, restaurants, and bars, for example, in the media and middle-up to lobby politicians against the proposal. Nevertheless, when Liverpool and London proposed passing local smoke-free legislation, when national legislation was not forthcoming, this cleverly separated the interests of the hospitality industry and the tobacco industry: the hospitality trade viewed national legislation as preferable to local laws and the ‘uneven playing field’ that would result.

ASH had only around five to seven members of staff during this period but enabled a multiplicative effect for the volume of advocacy, successfully scaling up its reach and effectiveness. For example, over 50% of the public were aware of the existence of ASH, and 92% of stakeholders rated ASH’s campaigning and policy work as excellent or good.^24^

### Case study 2. Increasing signalised crossing times for pedestrians in the UK

Using nationally-representative Health Survey for England data, Asher et al. demonstrated that 76% of men and 85% of women aged 65+ who could walk 8m unaided walked slower than the 1.2m/s threshold speed. Mean walking speed was 0.9m/s for men and 0.8m/s for women.^22^ On publication, there was massive traditional and social media coverage (https://academic.oup.com/ageing/article/41/5/573/47590#405680, https://oxfordjournals.altmetric.com/details/791287), noticed by non-governmental (civil society) organisations (NGOs) and community groups.

Stimulated by this media coverage, Living Streets, an NGO that promotes walking and campaigns for better conditions for pedestrians launched *‘3 Seconds More’* in November 2013 (https://www.livingstreets.org.uk/policy-and-resources/our-policy/crossings). This campaign aimed to increase the time available to cross an average road through reducing the assumed walking speed to 0.8m/s. Opposition came from motorists’ organisations and traditional transport planning approaches that prioritise motor vehicles, valuing car occupants’ time more than other road users’.

#### Middle-out activities

As the signalised crossings’ timings are mandated by national government, top actors were the Secretary of State (Minister) for Transport and the Minister for Roads, plus senior civil servants in the Department (Ministry) for Transport (DfT). They were the only actors with sufficient power to enable change.

The bottom actors in this arena were members of the public (particularly the elderly and those concerned for people with mobility impairments), local community groups, and individual practitioners who were members of professional organisations.

Middle actors were organisations interested in population health, transport, ageing, and/or inequalities, including local government and other practitioners’ professional associations; NGOs; and the media. One of the paper’s authors (JM) worked with Living Streets to include a broader set of middle actors, including the Transport and Health Study Group (an association of practitioners, policy-makers and researchers interested in implementing evidence-based policies to improve health and reduce inequalities associated with transport) and the UK Faculty of Public Health (the professional association for public health specialists).

Living Streets’ actions are summarised in Table 2, classified by the type and direction of action. More than 10,000 people wrote to their MP to support the campaign, asking the MPs to lobby the Secretary of State to give pedestrians 3 more seconds at signalised pedestrian crossings. The aggregation of actors’ voices and mediation between the levels increased the visibility of the crossing times issue, raised decision-makers’ awareness, and put it on their agenda.

**Table 2.** Increasing signalised crossing times for pedestrians in the UK.

| **Direction of action** | **Enabling<sup>a</sup>** | **Mediating<sup>a</sup>** | **Aggregating<sup>a</sup>** |
| --- | --- | --- | --- |
| Middle-Down | *Providing information:* Living Streets enhanced their supporters' capacity to act and engage by providing encouragement and guidance on how to raise this with their MP or DfT. | *Communication with the public:* Living Streets maintained communication with the public (via their own supporters). Other middle actors (professional associations, NGOs) also communicated with the public directly, using the media, or via their members/supporters. | *Formalisation of a grassroots initiative:* Living Streets contacted their supporters, who already had high motivation to engage with this issue but increased that by encouraging them to lobby the DfT directly or via their own MP. |
| Middle-Up | *Influencing the drafting of legislation:* As a result of their campaigning, based on the | *Translating between professional and sectoral languages:* Middle actors presented evidence (e.g. Asher et al., 2012, the TRL review) that fed in to the draft NICE *Guidance on* | *Aggregating the evidence:* Living Streets commissioned TRL to review the evidence on |
|  | Asher et al. paper<sup>22</sup> and the TRL report they commissioned, Living Streets staff were asked to review a chapter of DfT's updated *Traffic Signs Manual*. | *Physical Activity and the Built Environment*, responded to drafts, and lobbied the DfT and the relevant Ministers directly.<br>*Awareness-raising:* Middle actors used national media intensively to keep the issue live on decision-makers' agenda, e.g. publicising research by Living Streets of examples of individuals' difficulties in crossing the roads due to disabilities or poor crossing design (e.g. https://www.pressreader.com/uk/daily-mail/20170822/281676845029906 ) | crossing times and older people's walking speed, which confirmed Asher and colleagues' findings. |
| Sideways | *Providing information:* Living Streets brought the issue to other middle actors' attention and provided the evidence | *Building and maintaining coalitions with allies:* Living Streets involved the media and the other middle actors with good mediation capabilities, increasing their interest to | *Scaling up:* Living Streets invited other middle actors to encourage their members to join the advocacy efforts. |
|  | underpinning the problem for a substantial proportion of the population in trying to cross roads safely. | participate by bringing the issue to their attention and providing the evidence underpinning the problem. |  |

<sup>a</sup> Enabling: enabling action by using own resources and legitimacy to overcome barriers; Mediating: providing a communications channel;
Aggregating: providing resources, e.g. knowledge, funding.
MP: Member of Parliament; DfT: Department for Transport; NICE: National Institute for Health and Care Excellence.

One grassroots response to the media coverage was to create, perform and upload online a YouTube video *‘Hey Mr Boris’* by a campaigning choir of older people in a deprived area of London (https://www.youtube.com/watch?v=lpwboQxVJtg).

##### Outcomes

The *middle-up* impact was evident in May 2014, when the DfT announced consultation on *Traffic Signs Regulations and General Directions* (TSRGD), which includes crossings. DfT proposed that in future, Pelican crossings should not be installed – although existing crossings could remain. The strong support in the ensuing consultation of many middle actors, including several NGOs and two-thirds of local authorities, demonstrated the *middle-sideways* impact. In 2015, the DfT issued mandatory guidance that signalised pedestrian crossings installed in future must either provide a ‘countdown’ (Figure 2b) or be ‘Puffin crossings’(Figure 2c: these utilise a camera that keeps the lights green for pedestrians and red for other traffic while anyone is still walking across the junction).^25^

Further *middle-up* impact was evident in the National Institute for Health and Care Excellence *Guidance on Physical Activity and the Built Environment*, which recommended that local councils should ensure that pedestrian crossings allow adequate time for pedestrians to cross the road.^26^ In 2019, the DfT published updated guidance, permitting the use of a lower walking speed (1.0m/s) for signalised crossings where local authorities believe that will benefit local residents.^27^

## DISCUSSION

### Main findings of this study

ASH promoted smoke-free legislation, and Living Streets promoted change to crossing times, by acting as: mediators between the public interest and decision makers, and between various middle actors; aggregators, providing opportunities that amplified the voices of bottom and middle actors and made their demand more robust and visible; and enablers, proposing the evidence-based agenda as an alternative to the tobacco industry’s and car user lobbies’ agendas. Both NGOs increased the knowledge, interest and motivation of bottom and middle actors to actively engage in this domain and put pressure on decision-makers to act. They also increased the capacity of various, relatively small, diverse and widespread actors to act and present arguments to decision-makers at the top. The elevated motivation and capacity of top, bottom and middle actors facilitated the action.

Both case studies had as the ultimate aim changing national policy. In both, the main actions were sideways to multiply the effects of both middle-down and thus bottom-up, and of middle-up. Applying the MOP lens recognises the advocacy work that middle actors were uniquely positioned to lead in both these examples.

### What is already known

Learning from past public health campaigns can help in planning effective strategies for future campaigns. Involving key organisations and creating networks and alliances are important strategies for effective public health action ^28^. While such networks have commonly involved a wide range of health-relevant organisations and individuals,^29^ the adoption of a Sustainable Development Goal hygiene indicator on handwashing resulted from NGOs, academics, and commercial organisations working together with traditional public health bodies to influence policy-makers (middle-up), while implementation involves the same actors working middle-down.^30^ Many would argue that this, and the MOP, is how public health works, and has always worked.

### What this study adds

We suggest that using the MOP framework as a diagnostic lens and *formal* structure can assist public health professionals and others to identify the ‘missing middle actors’ and the interactions between them and other key actors. A more systematic approach would help in the design of advocacy or implementation strategies to achieve desired policy or behaviour changes and amplify the effectiveness of sideways, middle-up, middle-down, top-down and bottom-up activities.

The MOP can lead to public health practitioners stepping back and working in the background, leaving more overt action to others. While this low profile may be problematic for some individuals, or for justification of resources such as staff time, the goal should be the outcome in terms of the benefits for population health rather than the visibility of public health departments. Public health advocacy is a core skill of public health, yet the requisite skills and qualities are challenging, including familiarity with the evidence base and ability to effectively articulate key messages and relevant narratives to influence opinion leaders and the general public.^31^ The process can also involve potential conflicts in the blending of science, politics and activism in the context of wider public interest, such as the different timeframes of politicians and outcomes of effective public health measures,^32^ yet also has the power to deliver major systemic change. Legislation has a role reducing non-communicable diseases^32,33^; many recent public health laws that have been implemented were achieved through use of a middle-out approach, including banning tobacco marketing, plain packaging of tobacco, and nutrition labelling. In case study 1, the lead middle actor recruited a broad set of middle actors, including many who are not traditionally involved in public health work. In case study 2, most of the middle actors were more traditionally involved in promoting the health of the public. In both case studies, the lead actor was an NGO but that role may be taken by local government, public health bodies or departments, community groups, or others.

It should be recognised that those with opposing goals may also use a middle-out approach. For example, the tobacco industry involved the National Federation of Retail Newsagents and the Tobacco Retailers Alliance (membership organisations for newsagents and tobacconists) in opposing legislation to ban tobacco advertising^9^ and the hospitality industry to oppose proposed smoke-free legislation.^23^ Such efforts include apparent bottom-up activities using manufactured ‘grass-roots’ campaigns, referred to as ‘astroturfing’.^9^ Many health-diminishing industries have used techniques trialled by the tobacco industry^34^; proponents of good health can also learn lessons.^9,35^ The tobacco industry formerly, and the food and beverage industry more recently, have used a ‘sideways’ approach, involving national and international sporting bodies and individual clubs to promote unhealthy products to those attending or watching such sporting events (‘middle-down’).^36^ It is not known whether these bodies also support their sponsors’ interests in a ‘middle-up’ fashion. Thus, despite valid concerns about engaging with directly health-diminishing industries,^34^ public health organisations need to engage with the potential industry middle actors nationally and locally to promote health.

The United Nations (UN) Inter-Agency Task Force on the Prevention and Control of Non-communicable Diseases has called for increasing effective health-promoting partnerships with civil society and the commercial sector, giving due regard to managing conflicts of interests. These include stronger regulation and legislation to provide an environment that enables behaviours that promote health.^37^ Much of this can be facilitated by taking a middle-out approach, which assists formal consideration of the broader range of organisations and groups that could be involved as allies. The MOP can also help with the systems thinking that is now recognised as crucial in improving population health.^10^

## Limitations of this study

The main limitation is that the two case studies may not be representative. They were selected because we believe they illustrate the impact of middle actors. The MOP may be more or less applicable to other public health issues.

## Data Availability

All data produced in the present work are contained in the manuscript.

## CRediT statement

Conceptualisation: JS Mindell, Y Parag

*Methodology*: KB Janda, Y Parag

*Writing*: Original Draft: JS Mindell, Y Parag, S Bartington

*Writing:* Review & Editing: J Barlow, S Bartington, JS Mindell, Y Parag, L Stoll, KB Janda

*Visualisation:* Y Parag, JS Mindell

*Supervision:* JS Mindell, Y Parag

*Project administration:* I Hamilton, KB Janda, Y Parag, JS Mindell

*Funding acquisition:* KB Janda, Y Parag, I Hamilton

## Acknowledgements

We thank all participants in the Middle-Out Perspectives workshop held in Herzliya, Israel, 11-12 March 2020, for their insights. We also thank the workshop funders. We thank Action on Smoking and Health and Living Streets for their leadership in many public health campaigns, such as the two described as case studies in this paper.

